# Mapping Data Sources for Local Decision-Making on Maternal and Child Health in Tribal Primary Health Centre Settings

**DOI:** 10.64898/2026.03.28.26349587

**Authors:** Arun Mitra, Gurukartick Jayaraman, Bhavana Ondopu, Santosh Kumar Malisetty, Raghu Niranjan, Shaheen Shaik, Biju Soman, Rakhal Gaitonde, Tarun Bhatnagar, Engelbert Niehaus, K.S Sajinkumar, Adrija Roy

## Abstract

**Background:** Health systems in low- and middle-income countries are frequently described as “data rich, information poor,” collecting substantial data that rarely informs local decision-making. In tribal settings, this challenge is compounded by geographic isolation, fragmented governance, and the absence of disaggregated tribal health data within routine systems. We conducted a systematic mapping of data sources available for maternal and child health (MCH) decision-making at tribal Primary Health Centres (PHCs) in Andhra Pradesh, India.

**Methods:** Using a participatory data discovery approach embedded within an action research project, we mapped data sources across three PHCs under the Integrated Tribal Development Agency (ITDA) Rampachodavaram, Alluri Sitarama Raju District. Data discovery proceeded through three phases: document review, key informant interviews with Medical Officers and frontline health workers, and stakeholder validation. Sources were classified using the HEALTHY framework (Healthcare, Education, Access, Labour, Transportation, Housing, Income) and the Keller data discovery typology (Designed, Administrative, Opportunity, Procedural). Accessibility was assessed based on whether Medical Officers could retrieve data for local planning.

**Results:** We identified 28 distinct data sources relevant to MCH decision-making. Healthcare dominated (57.1%), while determinant domains remained underrepresented: Housing (10.7%), Income (10.7%), Education (7.1%), Labour (7.1%), Transportation (3.6%), and Access to healthy choices (3.6%). By data origin, Administrative sources predominated (46.4%), followed by Opportunity (21.4%), Procedural (17.9%), and Designed (14.3%). Despite 67.9% of sources having digital components, only 32.1% were fully accessible to Medical Officers, with 10.7% partially accessible and 57.1% inaccessible at the PHC level. Accessibility barriers were consistent across data categories, ranging from 50.0% to 66.7% inaccessibility.

**Conclusions:** The tribal PHC data ecosystem exhibits a fundamental mismatch between data generation and local utility. Data flows predominantly upward for administrative reporting rather than laterally for local decision-making. Addressing MCH outcomes in tribal populations requires reorienting health information systems toward local actionability.

## 1 Introduction

Health information systems in low- and middle-income countries (LMICs) have long been characterised by a paradox: vast quantities of data are collected, yet this data rarely translates into actionable information for local decision-making. Braa and Sahay termed this the “data rich, information poor” phenomenon, observing that health systems in developing countries invest heavily in data collection while failing to support data use at the point of care (1). This tension between data generation and data utilisation represents one of the most persistent challenges in global health systems strengthening (2).

In India, this paradox manifests acutely in tribal health settings. India’s 104 million Scheduled Tribe population experiences substantially worse health outcomes than national averages: under-five mortality stands at 50.3 per 1,000 live births compared to 41.9 nationally, institutional delivery reaches 82.3% versus 88.6%, and anaemia among tribal women has worsened to 64.6% compared to 57.0% nationally (3,4). Yet the Expert Committee on Tribal Health, in India’s first comprehensive assessment of tribal health, reported that “the darkness of information was astounding.” Routine health management information systems contain no mechanism to generate or track tribal-disaggregated data (5). If we cannot measure tribal health outcomes distinctly, we cannot effectively target interventions.

At the national level, India’s health data landscape has expanded substantially over the past decade. Mishra et al. documented 69 national healthcare data sources spanning routine service delivery, patient-based care, infrastructure, human resources, and financial data (6). The Health Management Information System (HMIS), launched in 2008, receives monthly reports from over 180,000 facilities, while the Reproductive and Child Health (RCH) Portal tracks individual beneficiaries with unique identifiers. Yet these national inventories do not assess what data reaches the local level for decision-making, nor do they capture the broader ecosystem of data relevant to health determinants in specific contexts.

The Primary Health Centre (PHC) represents the critical interface between India’s health system and its rural and tribal populations. Medical Officers at PHCs are expected to plan and implement maternal and child health programmes, identify high-risk cases, coordinate with frontline workers, and respond to local health challenges. Effective local planning requires access to data: not just on service delivery, but on population denominators, determinants of health, and contextual factors that shape health outcomes. In tribal areas administered by Integrated Tribal Development Agencies (ITDAs), governance is multi-departmental. Health, education, nutrition, livelihoods, and infrastructure fall under different departmental hierarchies, each maintaining separate data systems rarely integrated at the local level.

Despite the importance of local data ecosystems for health decision-making, no study has systematically mapped the full complement of data sources available to Medical Officers in tribal PHC settings in India. Existing assessments focus on national data infrastructure or evaluate specific systems (HMIS, RCH Portal) in isolation, without examining the broader ecosystem including non-health data relevant to health determinants.

This study addresses this gap by conducting a participatory inventory of data sources available for MCH decision-making at tribal PHCs in Andhra Pradesh. We apply two complementary classification frameworks: the HEALTHY classification from the 3-D Commission, which categorises data by health-relevant domain (Healthcare, Education, Access to healthy choices, Labour, Transportation, Housing, Income), and the data discovery typology from Keller et al., which classifies data by origin (Designed, Administrative, Opportunity, Procedural) (7,8). By combining content-based and origin-based classification, we aim to provide a comprehensive picture of what data exists, where it comes from, and whether it reaches the local level.

### 1.1 Study Objectives

#### Our objectives were to

1. Identify and enumerate data sources relevant to MCH available within the tribal PHC data ecosystem
2. Classify sources by HEALTHY domain and data discovery category
3. Assess accessibility of each source to Medical Officers for local planning
4. Characterise the format, frequency, and granularity of available data

## 2 Methods

### 2.1 Study Design and Setting

This study was conducted as the diagnostic phase of an action research project on participatory data science for maternal and child health in tribal settings. Following the classic action research cycle articulated by Susman and Evered, the project proceeds through iterative stages of diagnosing, action planning, action taking, evaluating, and specifying learning (9). Researchers and practitioners collaborate to generate knowledge through action within the system. The data source mapping reported here constitutes the “diagnosing” stage, establishing baseline conditions and identifying problems prior to collaborative intervention design with local stakeholders.

The study was conducted across three PHCs (Boduluru, Gangavaram, and Vadapalli) under ITDA Rampachodavaram in Alluri Sitarama Raju District, Andhra Pradesh, India. Alluri Sitarama Raju is a predominantly tribal district carved from East Godavari in 2022, with over 70% Scheduled Tribe population. The study PHCs serve populations ranging from approximately 15,000 to 25,000, comprising predominantly Koya and Konda Reddi tribal communities. The terrain is hilly with seasonal road accessibility constraints during monsoon months (June to September). Each PHC has one Medical Officer, two to three staff nurses, and coordinates with four to eight sub-centres staffed by Auxiliary Nurse Midwives (ANMs) and Accredited Social Health Activists (ASHAs).

### 2.2 Conceptual Framework

We employed two complementary classification frameworks to characterise the data ecosystem.

**The HEALTHY Classification** was developed by the Rockefeller Foundation and Boston University 3-D Commission to systematically categorise data sources relevant to social determinants of health (7,10). The seven domains are:

- **H**ealthcare: clinical services, health programmes, disease surveillance
- **E**ducation: schooling, literacy, health education
- **A**ccess to healthy choices: food security, water, sanitation, healthy environments
- **L**abour: employment, livelihoods, working conditions
- **T**ransportation: road access, connectivity, mobility
- **H**ousing: shelter quality, land tenure, settlement patterns
- **Y** (Income): economic status, financial inclusion, social protection

**The Keller Data Discovery Typology** classifies data sources by origin rather than content (8):

- **Designed data**: generated through deliberate scientific inquiry (surveys, clinical trials, special studies)
- **Administrative data**: generated through government or organisational processes (registrations, service records, reports)
- **Opportunity data**: generated passively through daily activities (mobile phone data, satellite imagery, sensor data)
- **Procedural data**: generated through institutional workflows and standard operating procedures (registers, stock records, meeting minutes)

This dual classification allows us to identify both thematic gaps (which HEALTHY domains are underrepresented) and structural gaps (which types of data are accessible).

### 2.3 Data Collection Process

Data source discovery proceeded through three iterative phases between October 2023 and March 2024.

#### Phase 1: Document Review (October to November 2023)

We systematically reviewed official documentation including: (a) registers maintained at PHC and sub-centre levels; (b) reporting formats submitted to ITDA, District Medical and Health Office, and state health directorate; (c) national programme guidelines for MCH services (RCH, immunisation, nutrition); (d) ITDA administrative records and departmental coordination mechanisms; and (e) digital system documentation (HMIS, RCH Portal, ANMOL, E-ASHA). Document review identified formal data sources within the health system and adjacent departmental systems.

#### Phase 2: Key Informant Interviews (December 2023 to February 2024)

We conducted semi-structured interviews with Medical Officers (n=3), staff nurses (n=4), ANMs (n=6), ASHAs (n=4), and ITDA health coordinators (n=2). Interviews explored: (a) what data sources participants used in daily practice; (b) what data they wished they had access to; (c) perceived barriers to data access; and (d) informal data sources and workarounds. Interviews were conducted in Telugu, the local language, and lasted 30 to 60 minutes. In addition, insights from Anganwadi workers were obtained through weekly convergence meetings held every Thursday at each PHC, where ANMs and Anganwadi workers jointly review beneficiary data and resolve discrepancies in MCH records. This phase revealed data sources not captured in formal documentation, including informal information practices and cross-departmental data.

#### Phase 3: Stakeholder Validation (March 2024)

Preliminary findings were presented at a validation workshop with PHC staff from all three facilities. Participants reviewed the inventory for completeness, corrected classifications, and verified accessibility assessments. This participatory validation ensured the inventory reflected local realities rather than researcher assumptions.

### 2.4 Data Source Characterisation

For each identified data source, we extracted: (a) name and description; (b) generating agency/department; (c) HEALTHY domain(s); (d) Keller data discovery category; (e) format (digital, paper-based, or both); (f) frequency of data generation/update; (g) spatial granularity (individual, household, facility, administrative area); (h) whether spatial coordinates were available; and (i) accessibility to Medical Officers for local planning.

#### Accessibility was assessed through a three-level classification

- **Accessible (Yes):** Medical Officer can directly retrieve or has routine access to data
- **Partially Accessible:** Medical Officer can request data but faces barriers (approval requirements, delays, format incompatibility)
- **Inaccessible (No):** Data exists but Medical Officer cannot access for local use

We distinguished “Traditional” sources (those embedded in routine health information systems) from “Extended” sources (data from non-health departments or unconventional sources with relevance to health determinants).

### 2.5 Ethical Considerations

This study was approved by the Institutional Ethics Committee, Sree Chitra Tirunal Institute for Medical Sciences and Technology (Protocol No. SCT/IEC/2047/MAY/2023). Written informed consent was obtained from all interview participants. The study was conducted in accordance with the Declaration of Helsinki. No individual patient data was accessed; the study focused on data *sources* rather than data *content*.

## 3 Results

### 3.1 Overview of the Data Ecosystem

We identified 28 distinct data sources relevant to maternal and child health decision-making within the tribal PHC ecosystem (Table 1). Of these, 11 (39.3%) were Traditional sources embedded in routine health information systems, 14 (50.0%) were Extended sources from non-health departments or unconventional data streams, and 3 (10.7%) spanned both categories.

**Table 1.**
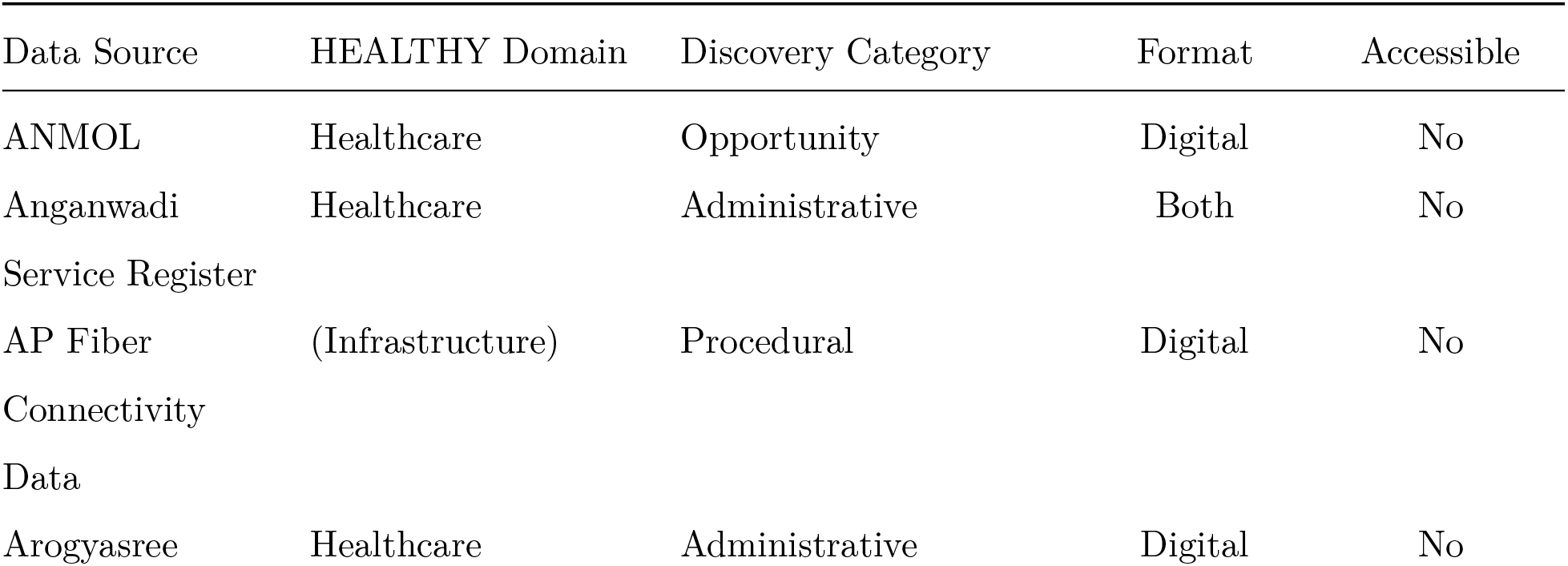
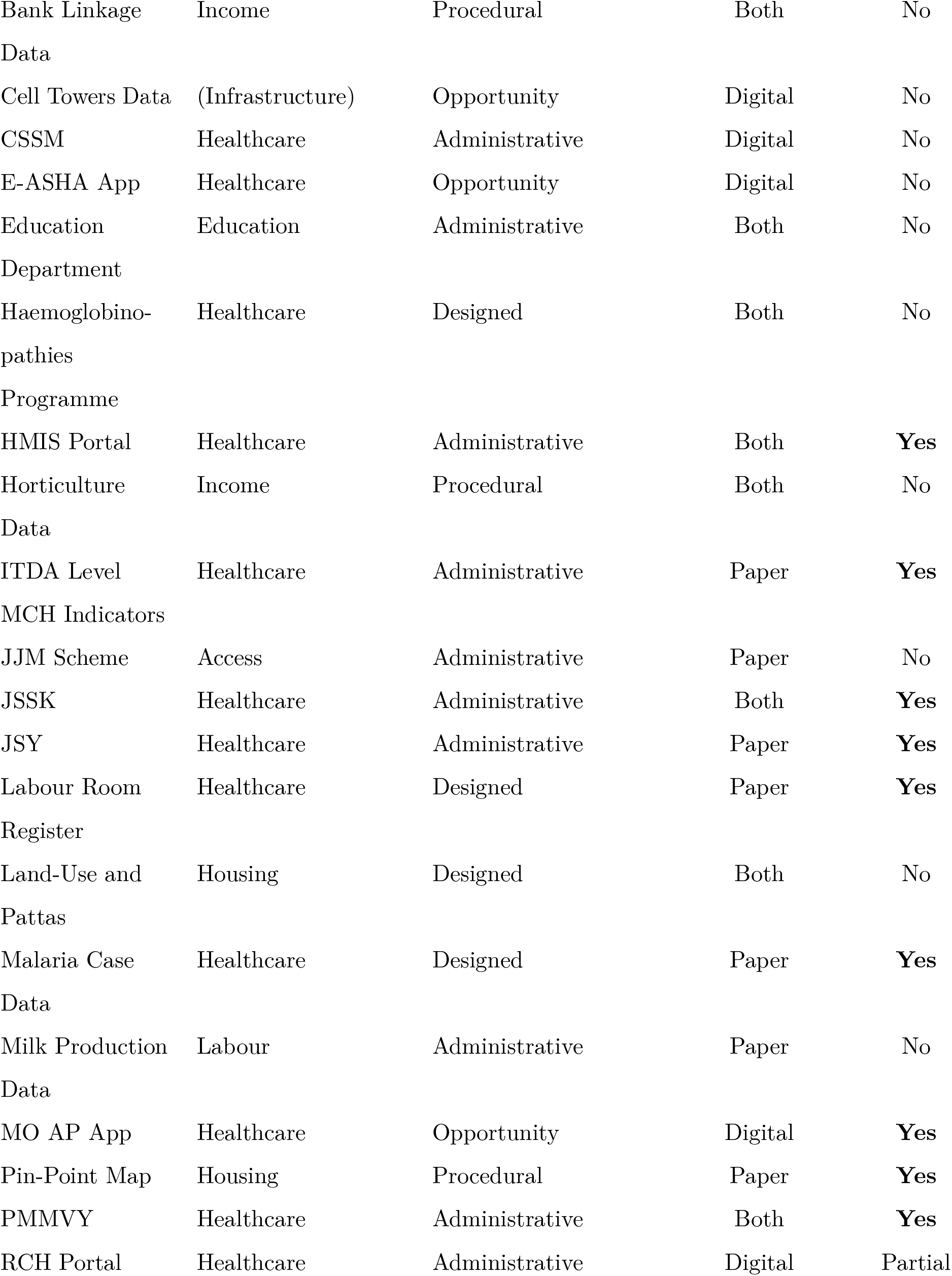
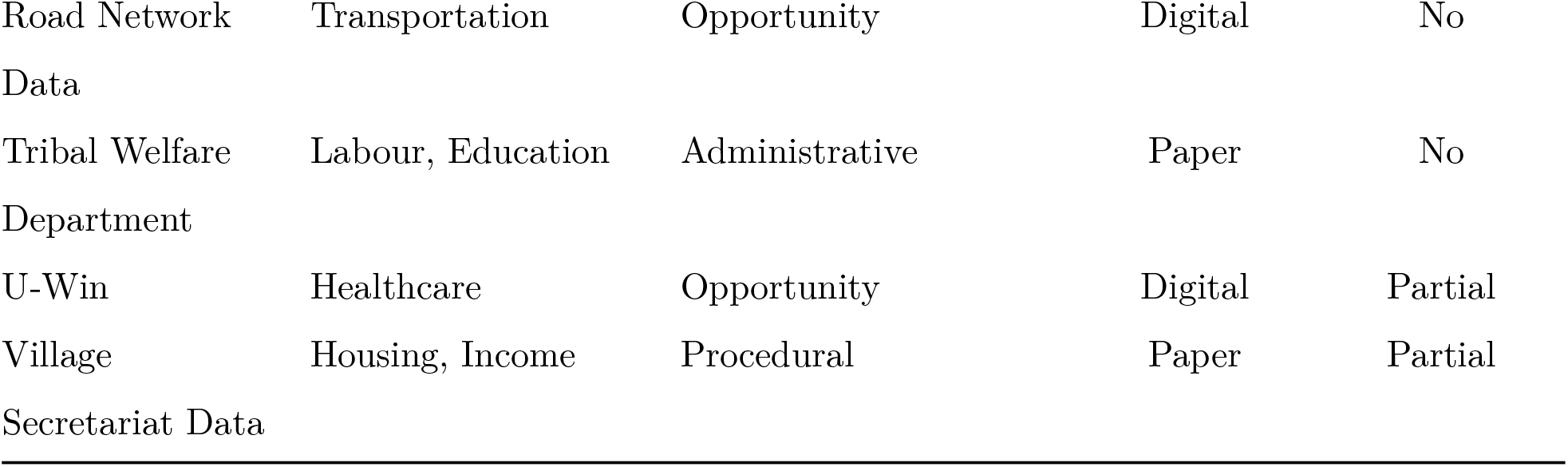
Data Sources Identified in the Tribal PHC Ecosystem (*n*=28) Full inventory of data sources with classification by HEALTHY domain, Keller category, format, and accessibility to Medical Officers. HMIS = Health Management Information System; RCH = Reproductive and Child Health; CSSM = Child Survival and Safe Motherhood; JSY = Janani Suraksha Yojana; JSSK = Janani Shishu Suraksha Karyakram; PMMVY = Pradhan Mantri Matru Vandana Yojana; ITDA = Integrated Tribal Development Agency; JJM = Jal Jeevan Mission.

### 3.2 Distribution by HEALTHY Domain

Healthcare dominated the data ecosystem, with 16 of 28 sources (57.1%) classified under the Healthcare domain (Figure 1). Social determinant domains were substantially underrepresented: Housing and Income each accounted for 3 sources (10.7%), Education and Labour for 2 sources each (7.1%), while Transportation and Access to healthy choices were each represented by only 1 source (3.6%). Two sources (AP Fiber Connectivity Data, Cell Towers Data) were not classifiable within HEALTHY domains as they represented infrastructure metadata rather than health-relevant content.

**Figure 1.**
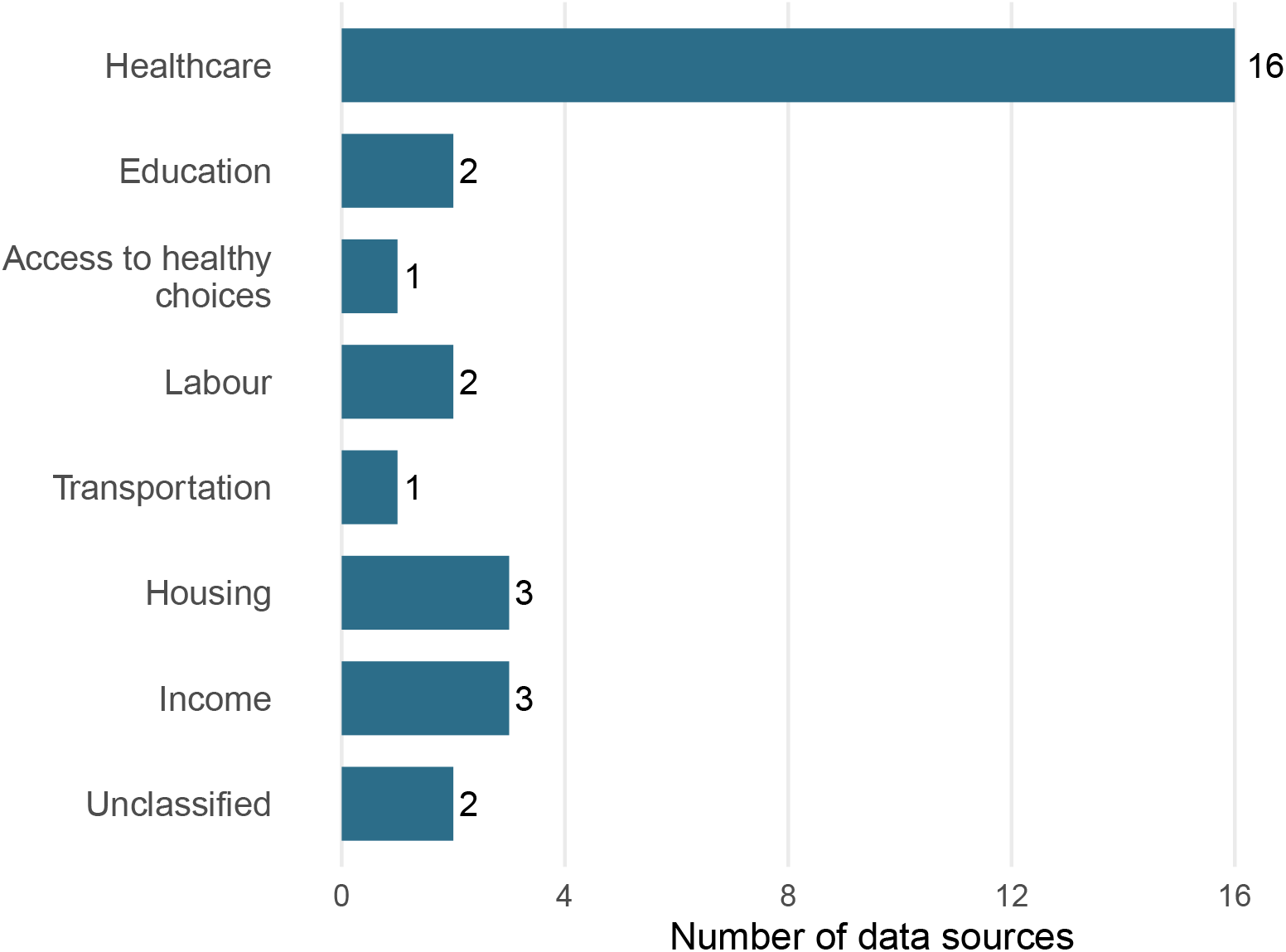
Distribution of 28 identified data sources across HEALTHY domains. Healthcare dominates (57.1%), while social determinant domains are underrepresented. Two sources (AP Fiber Connectivity Data, Cell Towers Data) were not classifiable within HEALTHY domains as they represent infrastructure metadata.

Healthcare sources included the major national reporting systems (HMIS Portal, RCH Portal, CSSM), maternal health schemes (JSY, JSSK, PMMVY), immunisation tracking (U-Win), mobile applications (ANMOL, E-ASHA, MO AP App), and facility-level registers (Labour Room Register, Malaria Case Data). The concentration in Healthcare reflects the health department’s primary mandate but leaves Medical Officers without systematic access to determinant data essential for understanding *why* MCH outcomes differ across populations.

### 3.3 Distribution by Data Discovery Category

Administrative data sources predominated, comprising 13 of 28 sources (46.4%) (Table 2). Opportunity data accounted for 6 sources (21.4%), Procedural data for 5 sources (17.9%), and Designed data for 4 sources (14.3%).

**Table 2.**
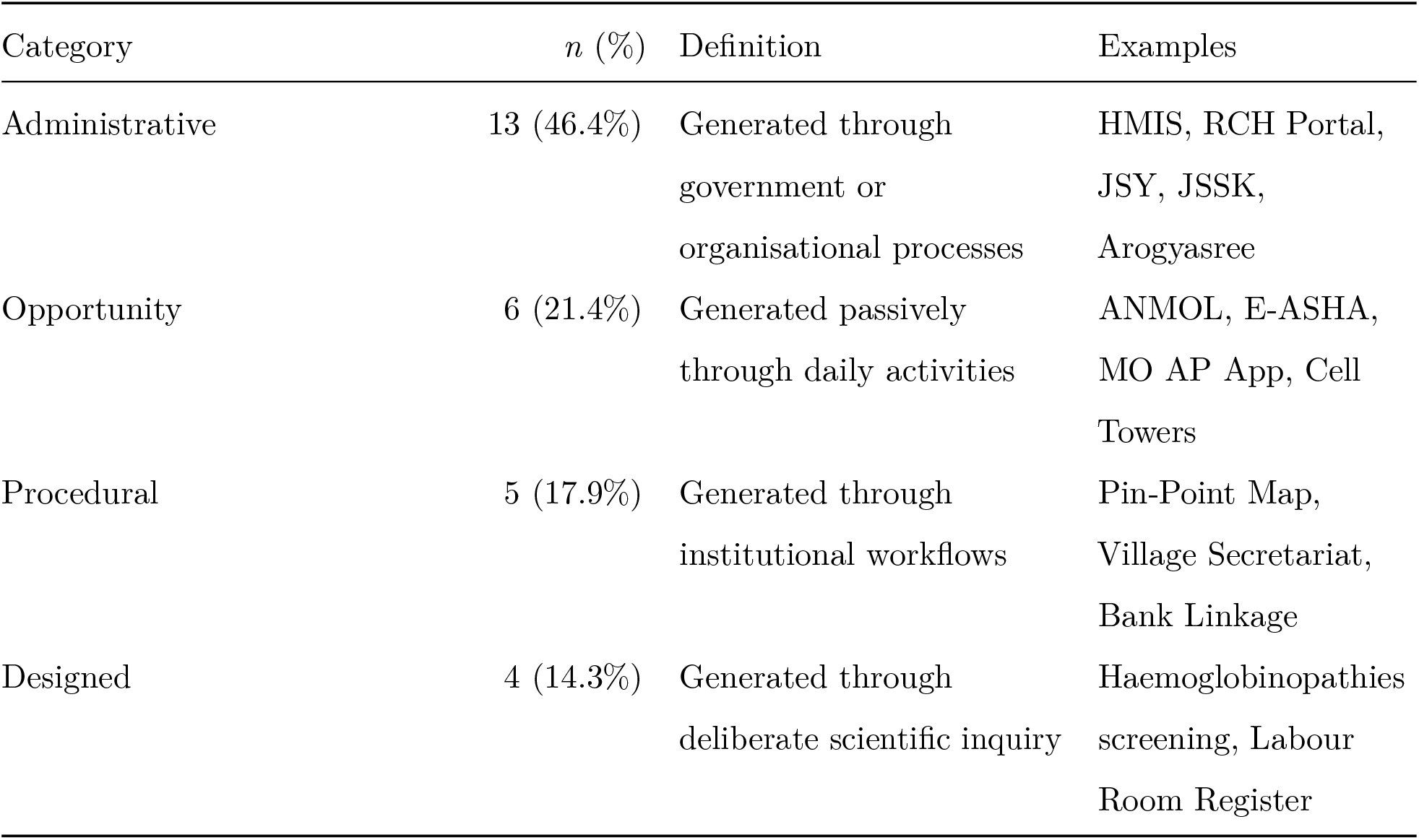
Distribution of Data Sources by Keller Data Discovery Category. Distribution of data sources by origin-based (Keller) classification. Category *n* (%) Definition Examples Administrative 13 (46.4%) Generated through government or organisational processes Opportunity 6 (21.4%) Generated passively through daily activities Procedural 5 (17.9%) Generated through institutional workflows Designed 4 (14.3%) Generated through deliberate scientific inquiry HMIS, RCH Portal, JSY, JSSK, Arogyasree ANMOL, E-ASHA, MO AP App, Cell Towers Pin-Point Map, Village Secretariat, Bank Linkage Haemoglobinopathies screening, Labour Room Register

| Category | *n* (%) | Definition | Examples |
| --- | --- | --- | --- |
| Administrative | 13 (46.4%) | Generated through government or organisational processes | HMIS, RCH Portal, JSY, JSSK, Arogyasree |
| Opportunity | 6 (21.4%) | Generated passively through daily activities | ANMOL, E-ASHA, MO AP App, Cell Towers |
| Procedural | 5 (17.9%) | Generated through institutional workflows | Pin-Point Map, Village Secretariat, Bank Linkage |
| Designed | 4 (14.3%) | Generated through deliberate scientific inquiry | Haemoglobinopathies screening, Labour Room Register |

Administrative sources included the major reporting systems (HMIS, RCH Portal, CSSM), scheme-based tracking (JSY, JSSK, PMMVY, Arogyasree), departmental records (Education Department, Tribal Welfare Department, Milk Production Data, JJM Scheme), and facility-level registers. These sources are generated through routine government operations and represent the backbone of formal information systems.

Opportunity sources included mobile applications generating data through field activities (ANMOL, E-ASHA, MO AP App, U-Win) and infrastructure data (Cell Towers, Road Network). These represent data produced as a byproduct of other activities that could be repurposed for health planning.

Procedural sources included institutional workflow documentation (Pin-Point Map, Village Secretariat data, Bank Linkage Data, Horticulture Data, AP Fiber Connectivity). These represent data generated through standard operating procedures that could inform health planning but were not designed for that purpose.

Designed sources were notably scarce (14.3%), comprising only the Haemoglobinopathies (Sickle Cell) screening programme, Labour Room Register, Land-Use/Pattas data, and Malaria Case Data. This reflects the absence of local survey capacity and the predominance of administrative over scientific data generation in the tribal PHC context.

### 3.4 Accessibility to Medical Officers

Of the 28 identified data sources, only 9 (32.1%) were fully accessible to Medical Officers for local planning (Table 3). Three sources (10.7%) were partially accessible, requiring requests or facing delays. Sixteen sources (57.1%) were inaccessible at the PHC level.

**Table 3.** Accessibility of Data Sources to Medical Officers by Category. Cross-tabulation of data discovery category and accessibility status.

| Discovery Category | Total | Accessible | Partial | Inaccessible | % Inaccessible |
| --- | --- | --- | --- | --- | --- |
| Designed | 4 | 2 | 0 | 2 | 50.0% |
| Administrative | 13 | 5 | 1 | 7 | 53.8% |
| Procedural | 5 | 1 | 1 | 3 | 60.0% |
| Opportunity | 6 | 1 | 1 | 4 | 66.7% |
| **Total** | **28** | **9** | **3** | **16** | **57.1%** |

Accessible sources included: HMIS Portal (with facility-level access), ITDA Level MCH Indicators (shared through monthly review meetings), JSSK and JSY records (maintained at PHC), Labour Room Register (PHC-maintained), Malaria Case Data (PHC-maintained), MO AP App (Medical Officer’s own device), Pin-Point Map (annual household enumeration), and PMMVY records.

Partially accessible sources were: RCH Portal (Medical Officer can view aggregate reports but cannot download individual-level data without district approval), U-Win (access limited to own facility’s immunisation records), and Village Secretariat data (can be requested through Gram Panchayat but not routinely available).

Inaccessible sources spanned both health and non-health systems. Within health systems, ANMOL, CSSM, and E-ASHA data are uploaded by ANMs and ASHAs but not viewable by Medical Officers. Arogyasree data flows directly to the insurance system without PHC access. Haemoglobinopathies screening data is maintained at district programme level. Non-health data (Education Department, Horticulture, Milk Production, Road Network, AP Fiber, Cell Towers, Land-Use, Tribal Welfare, JJM Scheme, Bank Linkage) is maintained by respective departments without mechanisms for cross-departmental sharing at PHC level.

Accessibility varied minimally by data discovery category: 50.0% of Designed sources were inaccessible, compared to 53.8% of Administrative, 60.0% of Procedural, and 66.7% of Opportunity sources. The similar inaccessibility rates across categories suggest that barriers are structural (relating to data architecture and governance) rather than category-specific.

### 3.5 Format and Technical Characteristics

The data ecosystem exhibited a mixed digital-paper landscape. Ten sources (35.7%) were digital-only, 9 (32.1%) were paper-only, and 9 (32.1%) existed in both formats (Figure 2). Digital presence did not ensure accessibility: of the 19 sources with digital components, only 4 (21.1%) were fully accessible to Medical Officers, with a further 2 (10.5%) partially accessible.

**Figure 2.**
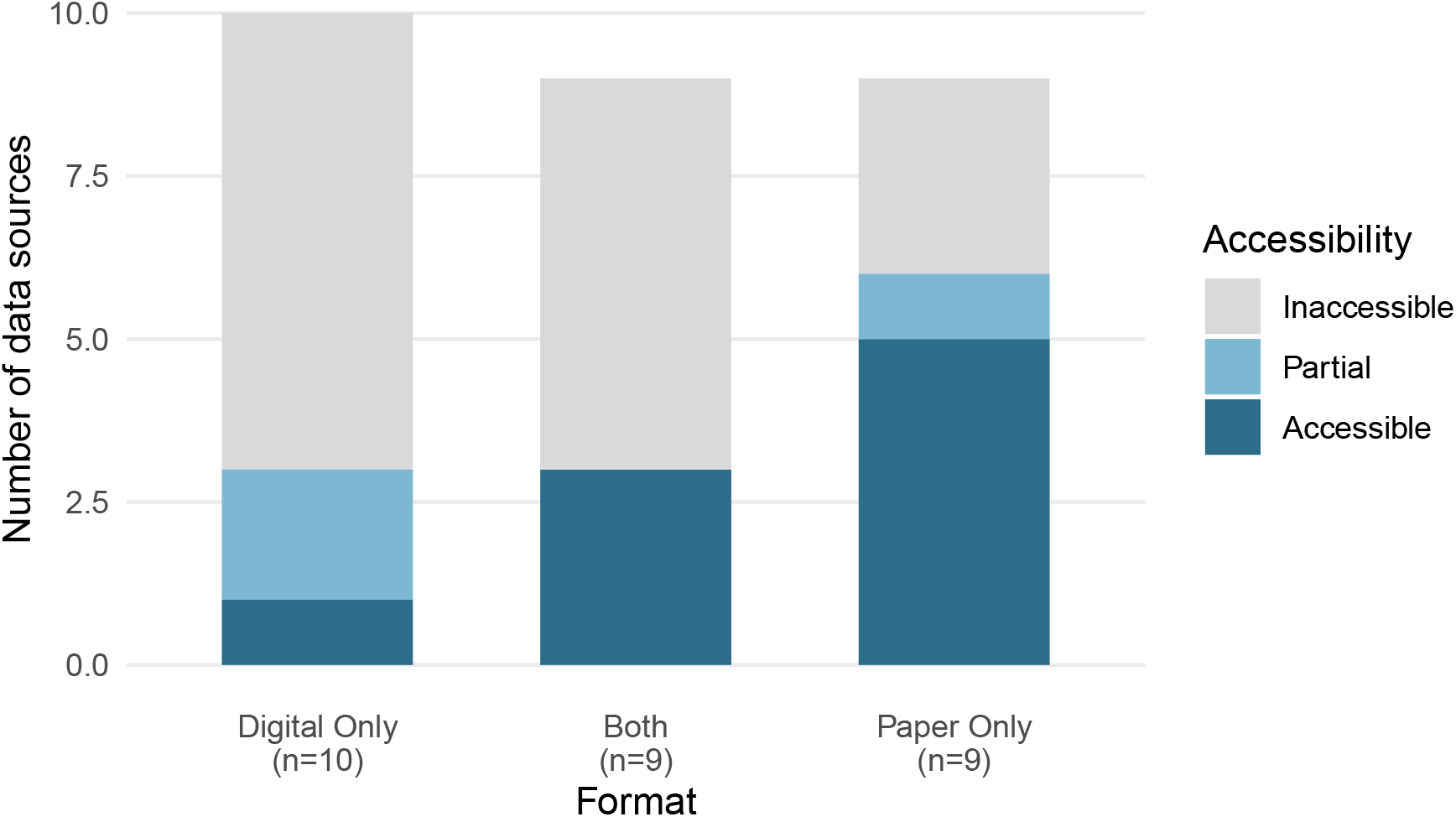
Format distribution and accessibility of data sources (n=28). Despite 67.9% of sources having digital components, only 32.1% were fully accessible to Medical Officers for local planning.

Temporal frequency varied substantially: 10 sources (35.7%) were updated monthly, 5 (17.9%) daily, 5 (17.9%) legacy/static, 4 (14.3%) yearly, with the remainder quarterly, weekly, or mixed frequency. Monthly administrative reporting cycles dominated, aligning with HMIS and scheme reporting requirements.

Spatial data was notably scarce: only 4 sources (14.3%) contained geographic coordinates or spatial identifiers usable for local mapping (Cell Towers, Land-Use/Pattas, Pin-Point Map, Road Network). This limits the capacity for spatial analysis and map-based decision support despite the geographic dispersion of tribal populations across difficult terrain.

Granularity ranged from individual-level tracking (RCH Portal, scheme beneficiaries) through household (Pin-Point Map), facility (HMIS), provider (ANMOL, E-ASHA), to administrative area aggregations. The RCH Portal and ANMOL contain individual-level data, but this granularity is inaccessible to Medical Officers who see only aggregate reports.

### 3.6 Key Accessible Sources for MCH Decision-Making

#### Among the 9 fully accessible sources, five are directly relevant to core MCH functions

1. **HMIS Portal:** Facility-level aggregate data on ANC, deliveries, and immunisation, uploaded monthly. Medical Officers have view access to their facility’s submitted data.
2. **Labour Room Register:** Paper-based record of all PHC deliveries with clinical details. Maintained by staff nurse, accessible on-site.
3. **Pin-Point Map:** Annual household enumeration by village/habitation with age-sex population counts. Used as denominator for coverage calculations.
4. **JSSK Records:** Beneficiary lists for free entitlements. Paper-based at PHC.
5. **Malaria Case Data:** Individual-level case register including pregnant women. Paper-based at PHC.

Notably, the individual-level data in RCH Portal that would enable case-based tracking is inaccessible to Medical Officers, leaving them reliant on aggregate reports that cannot identify specific high-risk beneficiaries.

## 4 Discussion

### 4.1 Principal Findings

This participatory inventory identified 28 data sources within the tribal PHC ecosystem relevant to maternal and child health decision-making. Three principal findings emerge.

First, the data ecosystem is healthcare-centric with limited determinant coverage. Healthcare sources accounted for 57.1% of identified data, while critical determinant domains were severely underrepresented. Transportation (3.6%) and Access to healthy choices (3.6%) were particularly sparse. In tribal settings where road accessibility, food security, and seasonal migration profoundly shape health-seeking behaviour and outcomes, this imbalance limits the capacity for comprehensive local planning. The ITDA governance structure coordinates health, education, livelihoods, and infrastructure, but data integration across these domains does not occur at the PHC level.

Second, accessibility does not follow digitisation. Despite 67.9% of sources having digital components, only 32.1% were accessible to Medical Officers. Digital systems designed for centralised monitoring create data that flows upward for administrative reporting but is not architected for lateral use at the facility level. ANMOL, for instance, enables ANMs to enter individual beneficiary data daily, yet Medical Officers cannot view this data. It is visible only to block and district supervisors. This architecture embodies what Sahay et al. termed “administrative control rationality” dominating over “care-based rationality” (11).

Third, designed data is scarce (14.3%), reflecting the absence of local survey or research capacity. Medical Officers lack mechanisms to generate data addressing local questions. The predominance of administrative data (46.4%) means available information reflects programme requirements rather than local information needs.

### 4.2 Comparison with National Data Landscape

Our findings complement and extend the national mapping by Mishra et al., who identified 69 healthcare data sources at the national level (6). While that inventory catalogued *what exists* nationally, our local-level assessment reveals *what reaches* the point of care. The national infrastructure (HMIS, RCH Portal, programme-specific systems) generates substantial data, but PHC-level accessibility is restricted.

The 57.1% inaccessibility rate we documented aligns with broader evidence on health information system challenges in India. Faujdar et al. found that among India’s major public health information systems, only DHIS2 supported local data use (12). NHM-HMIS was assessed as having “No” local data use functionality. Our findings provide granular confirmation that this architectural limitation translates into practical inaccessibility for frontline clinicians.

### 4.3 The “Data Rich, Information Poor” Paradox in Tribal Settings

Our findings provide empirical grounding for the “data rich, information poor” characterisation that Braa and colleagues advanced based on work across developing countries (1,2). Data *generation* is substantial: 28 distinct sources across multiple departments produce data relevant to MCH. But data *use* is constrained: only 9 sources reach Medical Officers in usable form.

The tribal PHC setting compounds this paradox through several mechanisms. Multi-departmental governance under the ITDA creates parallel data systems without integration mechanisms. Geographic isolation and connectivity constraints limit digital system access in remote sub-centres. The absence of tribal disaggregation in routine reporting renders tribal populations invisible within aggregate statistics. This confirms the Expert Committee on Tribal Health’s observation that institutional mechanisms to generate tribal-specific data “did not exist or did not function” (5).

### 4.4 Implications for Local Decision-Making

The data accessibility patterns we documented constrain local MCH planning in specific ways. Medical Officers cannot:

- **Identify high-risk beneficiaries** by name, because individual-level RCH Portal data is inaccessible
- **Map service coverage spatially**, because only 14.3% of sources contain spatial data
- **Integrate determinant information** (livelihoods, migration, food security), because cross-departmental data does not reach PHC level
- **Generate local evidence**, because designed data collection capacity is absent

These constraints limit the PHC’s capacity to function as envisioned in India’s health system architecture: as a planning and coordinating unit rather than merely a reporting node.

The PRISM framework identifies technical, organisational, and behavioural determinants of routine health information system performance (13). Our findings highlight technical determinants (system architecture that restricts data flow) and organisational determinants (multi-departmental governance without integration) that constrain data use regardless of individual motivation or skill. Addressing the “data rich, information poor” paradox requires architectural changes, not merely training or exhortation to use data.

### 4.5 Methodological Contributions

This study demonstrates the value of participatory data discovery within an action research framework. The three-phase approach (document review, key informant interviews, stakeholder validation) identified sources that would be missed by reviewing formal documentation alone. Informal practices, cross-departmental data, and tacit knowledge about data workarounds emerged through interviews with frontline workers who navigate these systems daily. The collaborative nature of action research ensured that the inventory reflected practitioner knowledge rather than researcher assumptions alone (9).

The dual classification using HEALTHY and Keller frameworks provides complementary perspectives. HEALTHY reveals *thematic* gaps (which determinant domains lack data), while Keller reveals *structural* characteristics (predominance of administrative over designed data). Together, they offer a comprehensive picture that single-framework approaches cannot provide.

This methodology is transferable to other settings seeking to understand local data ecosystems. The ODI Data Ecosystem Mapping methodology provides complementary visualisation tools (14). Future work could integrate inventory findings with ecosystem mapping to visualise actor relationships and data flows.

### 4.6 Recommendations

Based on our findings, we propose five recommendations for strengthening data availability for MCH decision-making in tribal PHC settings:

1. **Establish PHC-level data access** for individual-level RCH Portal data, enabling Medical Officers to identify and track high-risk beneficiaries rather than viewing only aggregate reports (15)
2. **Create cross-departmental data sharing mechanisms** at ITDA level, enabling integration of health, education, livelihoods, and infrastructure data for comprehensive local planning (16)
3. **Incorporate spatial data elements** into routine reporting, enabling map-based analysis of coverage and accessibility in geographically dispersed tribal populations (17)
4. **Design dashboard tools** that aggregate accessible data sources into actionable formats, building on existing accessible sources rather than requiring new data generation (15)
5. **Develop local data generation capacity** through simple designed data tools (rapid assessments, beneficiary feedback mechanisms) that address locally-relevant questions (16)

### 4.7 Limitations

This study has several limitations. First, the inventory was conducted in three PHCs under one ITDA; data ecosystems may differ in other tribal areas or states with different administrative arrangements. Second, we assessed accessibility from the Medical Officer’s perspective; ANMs and ASHAs may have different access patterns. Third, we documented data *sources* rather than data *quality*; accessible sources may contain incomplete or inaccurate data. Fourth, the classification of some sources across HEALTHY domains required judgment where content spanned multiple domains. Finally, the study period (October 2023 to March 2024) represents a snapshot. Data systems evolve, and new sources or access restrictions may have emerged subsequently.

### 4.8 Future Directions

This inventory provides the foundation for the subsequent action research cycles: co-designing a decision-support dashboard with PHC stakeholders that integrates accessible data sources for MCH planning (“action planning” and “action taking”), followed by evaluation and learning. The prioritisation of which data gaps to address (through advocacy for access, cross-departmental integration, or new data generation) will emerge through participatory processes with Medical Officers and frontline workers.

More broadly, systematic data source mapping should precede health information system interventions. Understanding *what data exists* and *what reaches local users* enables targeted interventions rather than generic capacity-building that may not address structural access barriers.

## 5 Conclusion

The tribal PHC data ecosystem in Andhra Pradesh contains 28 data sources relevant to maternal and child health, yet only 32.1% are accessible to Medical Officers for local decision-making. Healthcare data predominates (57.1%), while critical social determinant domains remain underrepresented. The majority of data flows upward for administrative reporting rather than serving local planning needs. Addressing MCH disparities in tribal populations requires reorienting health information systems from administrative monitoring toward local actionability, ensuring that the data generated about tribal communities serves those communities’ health.

## Data Availability

All data produced in the present study are available upon reasonable request to the authors

## Declarations

### Ethics approval and consent to participate

Approved by the Institutional Ethics Committee, Sree Chitra Tirunal Institute for Medical Sciences and Technology (Protocol No. SCT/IEC/2047/MAY/2023).

### Consent for publication

Not applicable.

### Availability of data and materials

The data source inventory is available as supplementary material.

### Authors’ contributions

AM conceptualised the study, conducted field data collection and analysis, and drafted the manuscript. GJ and AR contributed to field data collection. BO, SKM, RN, and SS, as PHC Medical Officers and action research co-participants, provided contextual insights throughout the research process and reviewed the draft. RG, TB, EN, and SK served on the doctoral advisory committee and contributed to conceptualisation, review of findings, and structuring of the draft. BS supervised the research and critically revised the manuscript. All authors approved the final version.

## Acknowledgements

We thank the ANMs, ASHAs, and ITDA staff who participated in this study and shared their knowledge of local data systems.

